# Effects of a feedback intervention on antibiotic prescription control in primary care institutions based on depth graph neural network technology: a cluster randomized cross-over controlled trial

**DOI:** 10.1101/2022.07.14.22277620

**Authors:** Junli Yang, Zhezhe Cui, Xingjiang Liao, Xun He, Shitao Yu, Wei Du, Shengyan Wu, Yue Chang

## Abstract

**Background:** Overuse and misuse of antibiotics are major factors in the development of antibiotic resistance in primary care institutions of rural China. In this study, the effectiveness of an artificial intelligence (AI)-based, automatic, and confidential antibiotic feedback intervention was evaluated to determine whether it could reduce antibiotic prescribing rates and avoid inappropriate prescribing behaviors by physicians.

**Methods:** A randomized, cross-over, cluster-controlled trial was conducted in 77 primary care institutions of Guizhou Province, China. All institutions were randomly divided into two groups and given either a 3-month intervention followed by a 3-month period without any intervention or vice versa. The intervention consisted of 3 feedback measures: a real-time warning pop-up message of inappropriate antibiotic prescriptions on the prescribing physician’s computer screen, a 10-day antibiotic prescription feedback, and distribution of educational brochures. The primary and secondary outcomes are the 10-day antibiotic prescription rate and 10-day inappropriate antibiotic prescription rate.

**Results:** There were 37 primary care institutions with 160 physicians in group 1 (intervention followed by control) and 40 primary care institutions with 168 physicians in group 2 (control followed by intervention). There were no significant differences in antibiotic prescription rates (32.1% vs 35.6%) and inappropriate antibiotic prescription rates (69.1% vs 72.0%) between the two groups at baseline (*p* = 0.085, *p* = 0.072). After 3 months (cross-over point), antibiotic prescription rates and inappropriate antibiotic prescription rates decreased significantly faster in group 1 (11.9% vs 12.3%, *p* < 0.001) compared to group 2 (4.5% vs 3.1%, *p* < 0.001). At the end point, the decreases in antibiotic prescription rates were significantly lower in group 1 compared to group 2 (2.6% vs 11.7%, *p* < 0.001). During the same period, the inappropriate antibiotic prescription rates decreased in group 2 (15.9%, *p* < 0.001) while the rates increased in group 1 (7.3%, *p* < 0.001). The characteristics of physicians did not significantly affect the rate of antibiotic or inappropriate antibiotic prescription rates.

**Conclusion:** The conclusion is that artificial intelligence based real-time pop-up of prescription inappropriate warning, the 10-day prescription information feedback intervention, and the distribution of educational brochures can effectively reduce the rate of antibiotic prescription and inappropriate rate.

**Trial registration:** ISRCTN, ID: ISRCTN13817256. Registered on 11 January 2020

## Introduction

Antibiotic resistance is a real threat to human health [1, 2]. Drug-resistant bacteria are continuously being discovered, and even some “superbugs” that are difficult to suppress with antibiotics have emerged [1]. In 2019, about 1.27 million deaths were related to antibiotic resistance [3]. Overuse and misuse of antibiotics are major factors in the development of antibiotic resistance [4]. The total consumption of antibiotics increased by 46% in 204 countries from 2000 to 2018 [5]. According to a World Health Organization (WHO) report the inappropriate use of antibiotics is on the rise, and is more likely to be found in low- and middle-income countries [6].

In China, more than 50% of outpatient antibiotic prescriptions are inappropriate [7] and this phenomenon is more prominent in primary care institutions [8]. In our previous retrospective investigation of 16 primary care institutions in Guizhou Province, approximately 90% of patients received inappropriate antibiotic treatment. The major inappropriate prescriptions were found in the patients diagnosed with diseases of the respiratory, digestive, and urinary systems [9] despite controlling for physicians’ individual prescribing behavior [10-14].

Previous researchers have developed a variety of interventions to control the misuse and overuse of antibiotic prescriptions, including information technology interventions, such as a Clinical Decision Support System (CDSS) or electronic health records, whereby electronic modules are sent to physicians to help them make the best clinical decisions [15-17]; educational interventions, such as distribution of educational brochures or training courses given to medical personnel or patients [18, 19]; and antibiotic prescription audit and feedback interventions [20-22]. We conducted a cluster randomized crossover-controlled trial based on a Hospital Information System (HIS) with 163 physicians in 31 primary care institutions in Guizhou Province in 2019. Significant results were achieved, with antibiotic prescription rates falling by 15% [23]. One limitation of the study, however, is that it did not take into account the rate of the rate of inappropriate antibiotics.

To this end, we introduce Depth Graph Neural Network technology (DGNN), an artificial intelligence (AI) deep learning algorithm. It is a new heterogeneous and complex network structure model and iterative optimization method [24]. The technology includes two parts: deep learning and graph neural networks. Deep learning is a type of machine learning technology based on representational learning of data; the technique mimics the way the human brain interprets data. Graph neural network is a deep learning model, which combines graph data with a neural network and performs end-to-end computation on the graph data. In recent years, DGNN has been used in medicine, organizational management, and marketing [25-27]. In view of this, based on prescription data and DGNN technology, a graph model of training data was established. At the same time, several shallow network structures were used to visualize the antibiotic use path graph and to realize the formulation and recommendation of an ideal treatment plan.

Therefore, an intelligent, confidential, and long-term feedback intervention warning system for inappropriate antibiotic prescription was thus developed. Among them, AI real-time warning was added on the basis of the previous feedback intervention study [23]. The objective of this study was to investigate whether the new feedback intervention could reduce the rates of antibiotic prescription and inappropriate prescription rates among primary care physicians.

## Methods design

### Trial designs and setting

A randomized, cross-over, cluster-controlled trial was conducted from April 1^st^, 2021 to September 30^th^, 2021. A cross-over design is a repeated measurement method in which each unit receives different interventions at different times [28]. In this study, a primary health care institution was used as a cluster unit. Physicians from the same institution were grouped together. As shown in Figure 1, all primary care institutions included in the trial were randomly divided into two groups: group 1 and group 2. The 3-month intervention was performed in group 1 while group 2 acted as the control group (no intervention given). As stated in the proposal [29], since this was a behavioral change intervention study, there was no washout period for this crossover design. Therefore, after 3 months, the two groups switched, with group 1 switching to be the control group and group 2 switching to receive the intervention for 3 months. The entire trial lasted for 6 months from April 1^st^, 2021 to September 30^th^, 2021 with the two groups entering the crossover point on June 30^th^.

**Figure 1.**
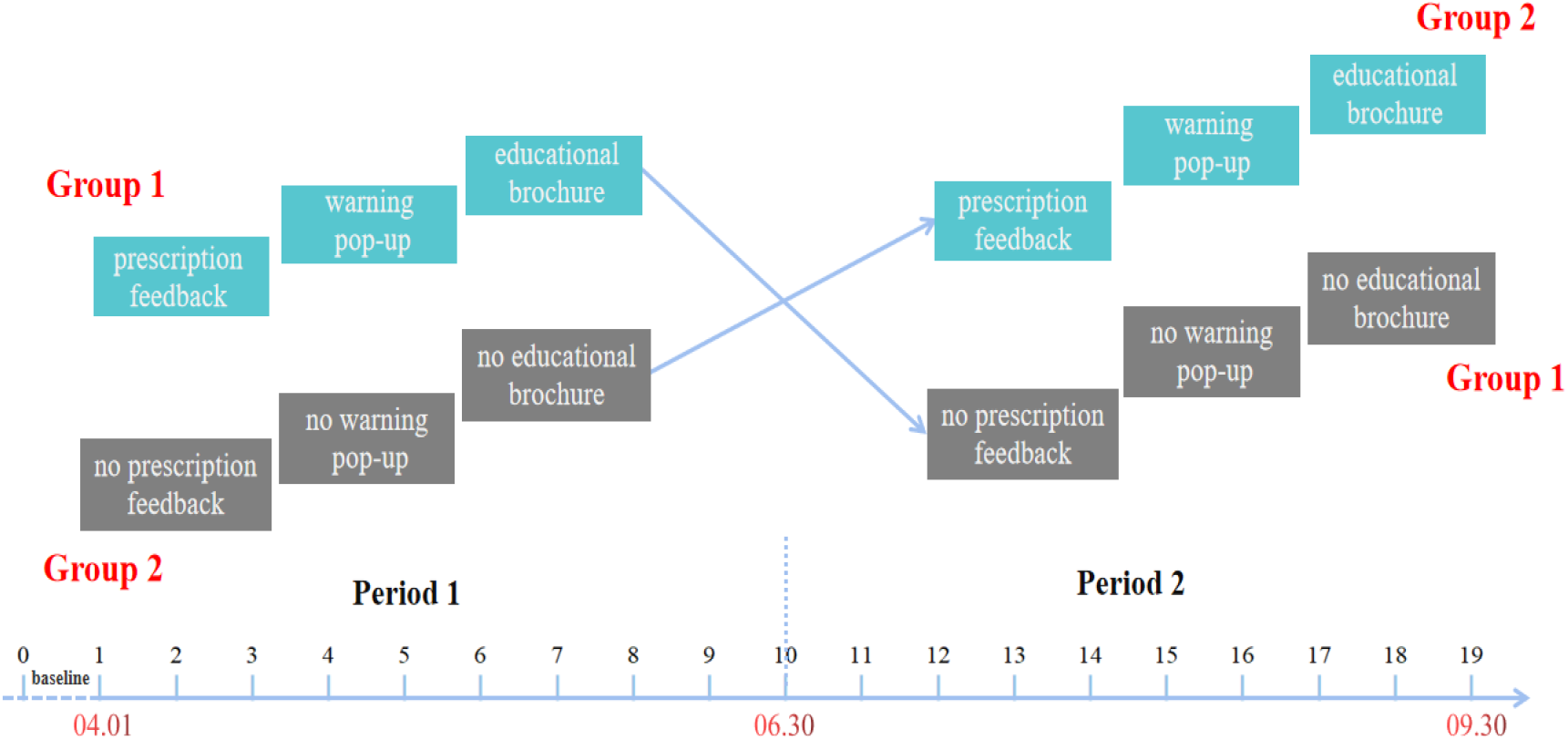
Cross-over intervention diagram showing 10-day intervals.

Guizhou Province, the setting of the study, is in the hinterland of southwest China and is one of the least developed provinces of the country. The study population involved 252 primary care institutions that use the same HIS in Guizhou Province. Township health centers and community health service centers are called primary care institutions, which mainly provide primary health care services for the local population [30]. The inclusion criteria were the same as in our previous study [23], and were also set out in the published protocol [29]: 1) institutions with at least 3 outpatient general physicians, 2) the physicians had worked in a primary care institution for at least one year, and 3) each physician saw at least 100 patients every 10 days. The exclusion criteria for prescriptions included patients treated for tuberculosis, leprosy and other diseases requiring combination drugs. Informed consent forms were signed by physicians before the trial commenced. One hundred thirty-two primary care institutions meeting the above criteria were included.

Antibiotic prescription records used in this trial were provided by Guizhou Lianke Weixin Technology Co., LTD. (LWTC). LWTC is a technology service company that develops and maintains medical and health information systems. Authorized by the Information Center Guizhou Provincial Health Commission (ICGPHC), an early warning intervention plug-in for antibiotic prescriptions was designed. The plug-in used DGNN technology to provide real-time warning and information feedback. Before the formal trial, the intervention plug-in had been successfully applied in two primary care institutions in Guizhou province for three months, and the sensitivity and reliability of the plug-in have been scientifically verified. The trial was approved by the Human Trial Ethics (Appendix 2) Committee of Guizhou Medical University (Certificate No.: 2019 (148)) in Dec. 27, 2019, and the protocol was published on January 7^th^, 2022 [29].

### Depth graph neural network technology (DGNN)

In the artificial intelligence (AI) part of this study, the representation of relevant data and knowledge for training and model evaluation was addressed. Specifically, based on the results of big data analysis, the influencing factors of physicians and patients on the rational use of antibiotics were summarized, and the Graph model-based knowledge representation and modeling method was studied in combination with the relevant contents of our self-made Guidance and Recommendations on Clinical Use of Antibiotics in Primary Care Institutions.

After solving the representation problem of Graph model of training data, the Depth Graph Neural Network (DGNN) technology with Directed Graphs structure and edge-informative Graph structure was studied. Specifically, a new heterogeneous and complex network structure model and iterative optimization method was used. The DGNN method made use of several shallow network structures at the same time, with the depth of the traditional neural network dozens or even hundreds of layers in the stack to achieve higher network expression ability and performance. It can effectively avoid the traditional deep learning technology update iteration complex problem.

In order to improve the interpretability of the developed DGNN method in the process of antibiotic abuse assessment and analysis, the graph data representing antibiotic use path was visualized by similarity measurement and clustering technology based on graph data. Exploratory retrieval and presentation of multiple analysis results were provided to improve the comprehensibility and clinical reference value of the results of antibiotic prescription evaluation in this study.

### Randomization and masking

The 79 primary care institutions that met the criteria were randomly selected from the 132 using a random number table by LWTC information technology staff. In total, 335 qualified outpatient physicians were enrolled in the intervention trial. Figure 2 shows the flow chart of the trial. Physicians participating in the trial were randomly assigned to the two groups. All physicians involved in the study had a good sense of whether they were entering the intervention, so it was impossible to blind the participants and the researchers.

**Figure 2.**
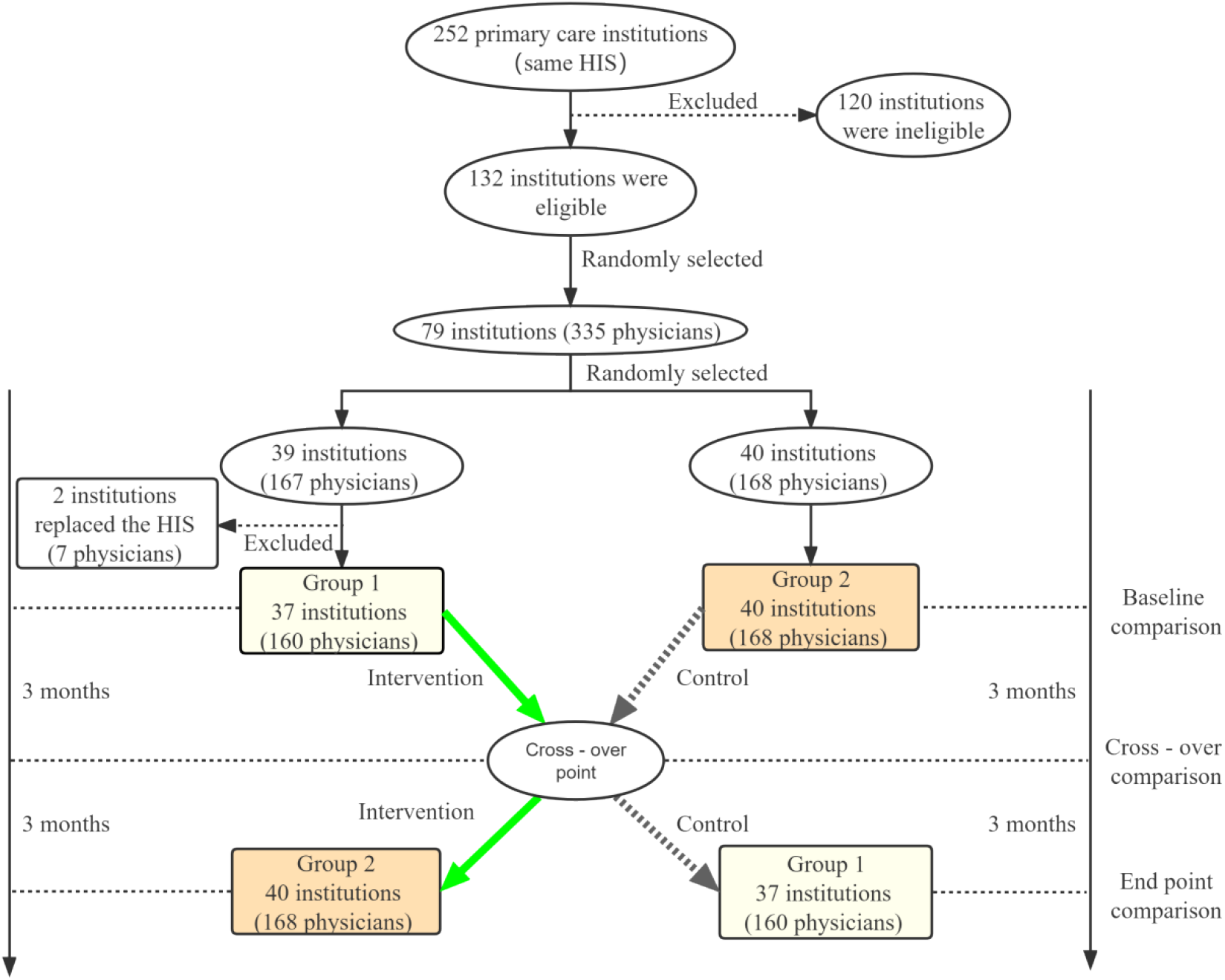
Flow chart of the trial.

### Intervention

Feedback interventions are the act of providing knowledge of the results of a behavior or performance to an individual [31, 32]. Feedback interventions can change behavior and improve performance and outcomes [16, 33]. Physicians who entered the intervention group received 3 feedback measures, including a real-time warning pop-up message, a 10-day antibiotic prescription feedback, and distribution of educational brochures.

The first measure of the feedback is the real-time warning of inappropriate antibiotics based on DGNN. In the HIS in China, when a physician prescribes an inappropriate antibiotic, a small window immediately appears reminding him of his inappropriate prescribing behavior. A brief explanation is also displayed in the message. There are 3 criteria for inappropriate antibiotic prescribing: 1) **unnecessary use**, such as patients who were diagnosed with viral infections but received antibiotics; 2) **incorrect antibiotic spectrum**, such as aminoglycosides are prescribed for gram-positive bacteria; 3) **combination of antibiotics without indication**, refers to the use of more than one systemic antibiotic in a visit, such as amoxicillin and levofloxacin in combination. Figure 3 shows an example of a pop-up message when an inappropriate antibiotic is prescribed. The feedback messages that the physicians received are actually shown in Chinese but here have been translated into English.

**Figure 3.**
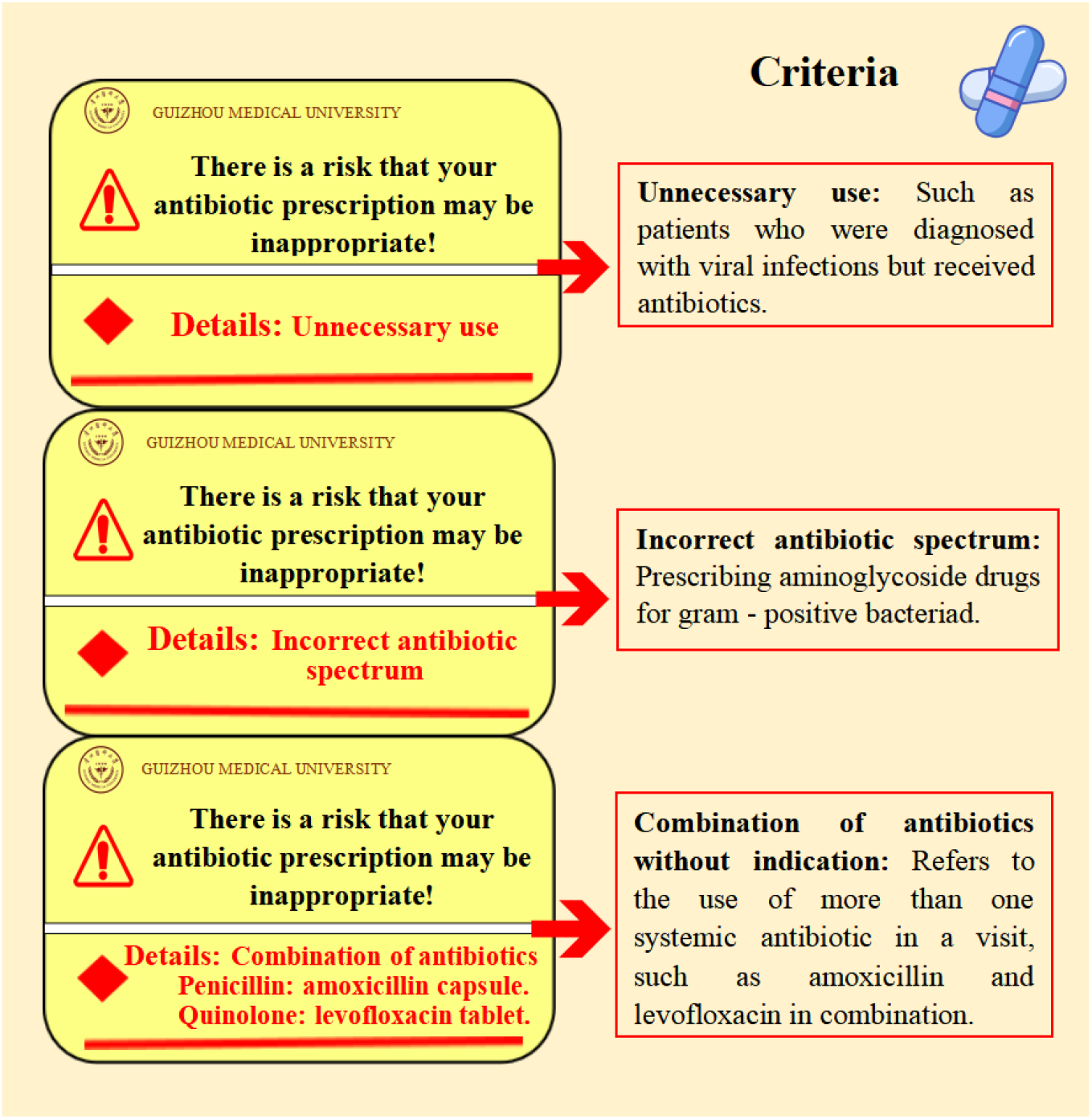
Example of real-time warning pop-up message.

The second measure of the feedback involves providing antibiotic prescription information to physicians every 10 days according to the HIS. The link to the prescription feedback information appears at the bottom of the physician’s computer screen, which the physician can click at any time and automatically updates every 10 days. As shown in Figure 4, the 10-day antibiotic prescription feedback included five functional areas. An automatic pop-up message was used to remind physicians to click on the link to view the information. The message was confidential; only the physician could see it. The physicians are free to pay attention to it or ignore it. The number of clicks per physician was also automatically recorded by the system.

**Figure 4.**
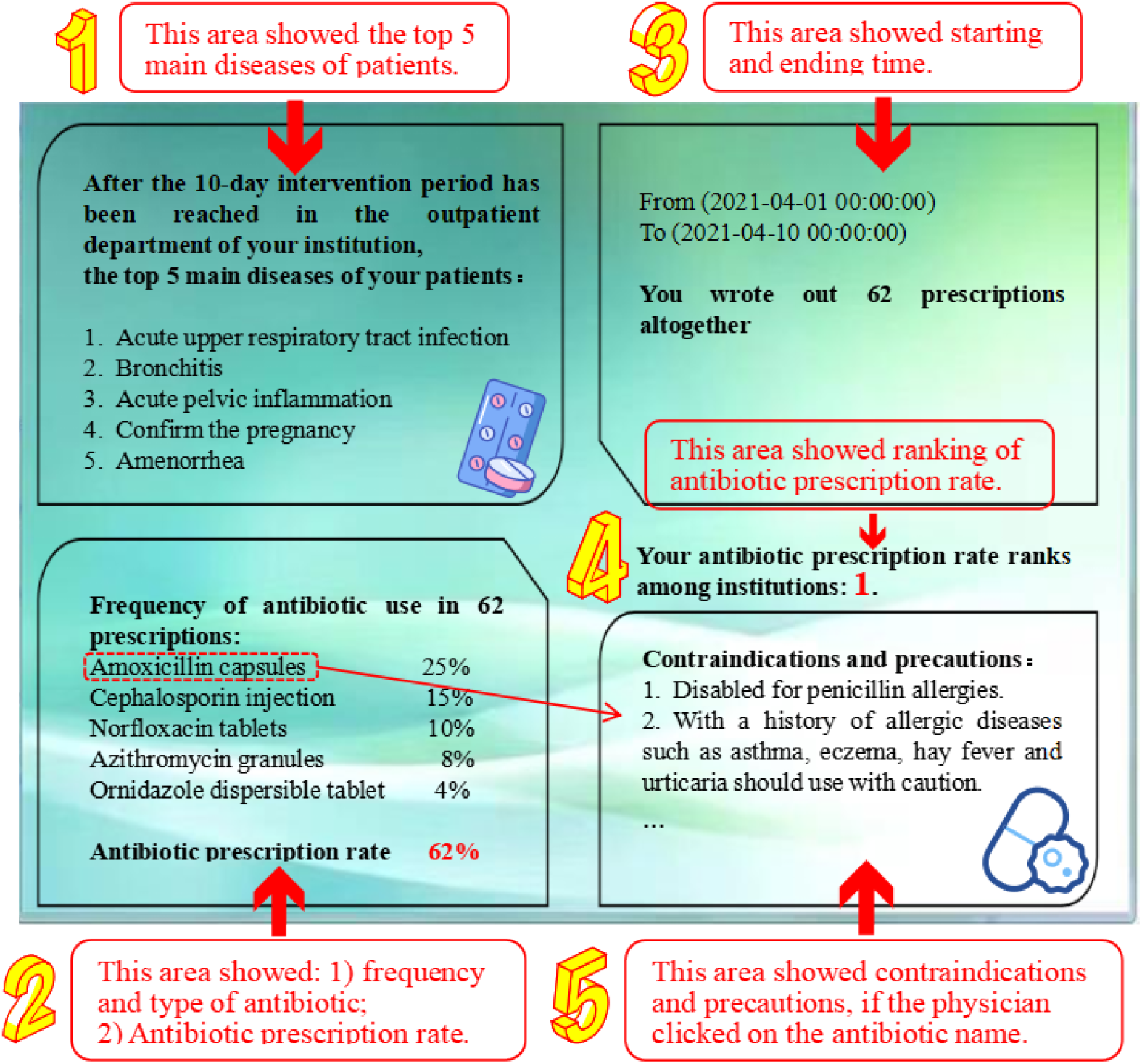
Example of a 10-day antibiotic prescription feedback.

The third part of the intervention is the educational brochures. Figure 5 shows a screen shot of the brochure’s cover, catalogue, and example of content (see Appendix 3 for details). The physicians could receive advice on antibiotic use and guidance on the diagnosis of common infections at the primary level. The number in the disease diagnosis section represents the diagnostic value, the higher the number, the higher the diagnostic value.

**Figure 5.**
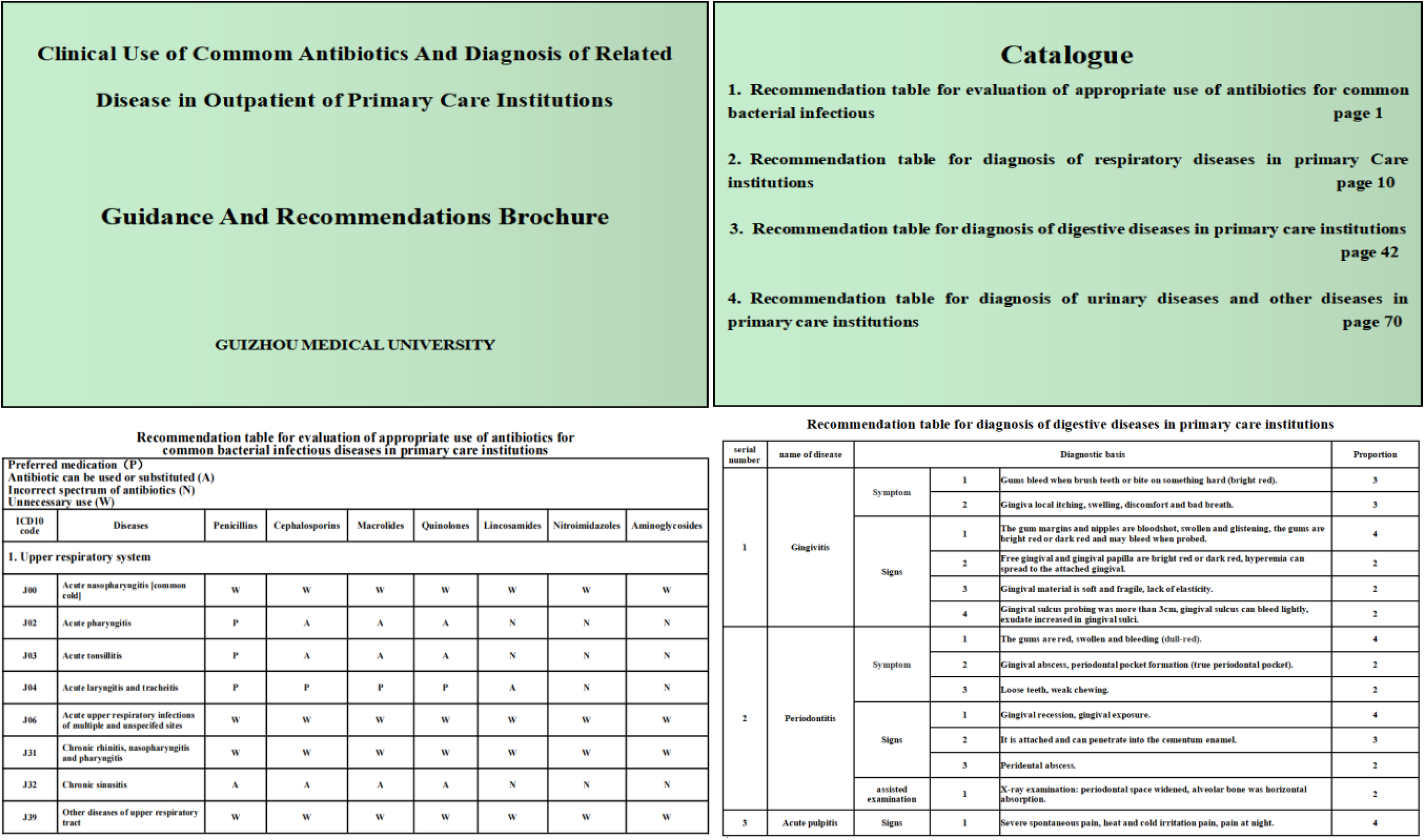
Example of an Educational Brochure.

### Control

No intervention was provided for the physicians in the control group who were advised to continue to treat patients as usual. During this period, all prescribing information from the physicians was recorded, but was not reported back to the physicians.

### Data collection and management

With the approval of ICGPHC, a data port was opened by LWTC technicians. We collected the antibiotic prescriptions, total prescription data, and relevant patient information from primary care institutions participating in the intervention trial. Codes were used to correlate the names of the physicians and patients in their prescriptions. Demographic information on physicians was obtained from the personnel department of the primary care institutions. All researchers involved in data collection signed confidentiality agreements.

The International Classification of Diseases, 10^th^ Edition (ICD-10) was used to classify the diseases of patients who were prescribed antibiotics. According to the list of essential medicines published by the World Health Organization, combined with the national guidelines for clinical application of antibiotics, the clinical application catalogue of antibiotics was summarized (Appendix 4) [34-36]. The antibiotics were categorized into seven classes, namely penicillins, cephalosporins, macrolides, quinolones, lincosamides, nitroimidazoles and aminoglycosides. Only systemic antibiotics were considered in this study; patients given external antibiotics such as erythromycin ointment and levofloxacin eye drops were excluded.

### Outcome variables

The primary outcome was the 10-day antibiotic prescription rate [23], which was the number of antibiotic prescriptions per 10 days divided by all prescriptions for 10 days. The secondary outcome was the 10-day inappropriate antibiotic prescription rate. The index was obtained by dividing the number of inappropriate antibiotic prescriptions by the total number of antibiotic prescriptions. An antibiotic prescription was determined to be inappropriate if any one of the following 3 criteria was satisfied: 1) unnecessary use; 2) incorrect antibiotic spectrum; 3) combination of antibiotics without indication. The characteristics of the physicians (age, sex, title, education, working years) and information related to antibiotics were included in the analysis as covariates.

### Sample size

The two independent means formula (two-tailed) was used to calculate the minimum number of physicians required for each group.

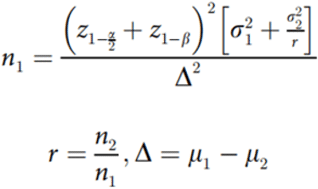

According to previous research experience [23], the average antibiotic prescription rate of the two groups were 35% and 30% and the variance was 15% for both. The type I error (*α*) was 0.05 and type II error (*β*) was 0.2. At least 142 physicians were needed in each group to observe the effects of the intervention. Considering a 10% attrition rate, at least 160 physicians were required in each group, for a total of 320.

### Statistical analyses

Student’s t test and rank-sum tests were used to compare the 10-day antibiotic prescription rate and the 10-day inappropriate antibiotic prescription rate between the two groups and within the same group. The effect of the intervention was measured by comparing antibiotic prescription rates and inappropriate rates at baseline, cross-over point, and end point. After the intervention, the antibiotic prescription rate and inappropriate prescription rate of physicians may not change immediately, and the magnitude of the change in rates may be different over time. Consequently, a transition model was used to predict the impact of the intervention on the antibiotic prescription rate and inappropriate antibiotic prescription rate. This model can adjust for statistical differences of physicians’ baseline characteristics before and after the cross-over point, and further explore the specific impact of the intervention on the antibiotic prescription rate. Spearman and Pearson correlation analyses were used to explore the relationship between antibiotic prescription rates and inappropriate antibiotic prescription rates during the intervention, as well as the relationship between physicians ranking and mouse-clicking frequency. Based on an intention-to-treat principle [37], data from outpatient physicians at all participating primary care institutions were included throughout the analysis (except for seven physicians at two hospitals where HIS was replaced). All data for this study was analyzed using R version 4.0.4.

## Results

A total of 79 primary care institutions consisting of 335 physicians were recruited. Thirty-nine primary care institutions containing 167 physicians were randomly assigned to group 1 (intervention followed by control) and 40 primary care institutions containing 168 physicians were randomly assigned to group 2 (control followed by intervention). However, in group 1, two primary care institutions were excluded because they had changed their HIS, so we were unable to obtain their prescription data (Figure 4). Overall, 313,165 antibiotic prescriptions were included in the analysis. Table 1 shows the baseline characteristics of physicians. The antibiotic prescription rates in group 1 and group 2 were 32.1% (10582/32938) and 35.6% (13097/36832), respectively. The two groups were similar in terms of sex, age, education, title and working years.

**Table 1.** Baseline characteristic of the physicians. [Mean ± SD or n (%)]

| Characteristic | group 1 (n = 160) | group 2 (n = 168) | Total (n = 328) |
| --- | --- | --- | --- |
| Antibiotic prescription rate | 32.1 | 35.6 | 33.9 |
| Sex |  |  |  |
| Male | 98 (61.2) | 108 (64.3) | 206 (62.8) |
| Female | 62 (38.8) | 60 (35.7) | 122 (37.2) |
| Age |  |  |  |
| 21-31 | 58 (36.3) | 61 (36.3) | 119 (36.3) |
| 32-41 | 44 (27.4) | 59 (35.1) | 103 (31.4) |
| 42-65 | 58 (36.3) | 48 (28.6) | 106 (32.3) |
| Education |  |  |  |
| Technical secondary school | 23 (14.3) | 27 (16.1) | 50 (15.2) |
| Junior college | 58 (36.3) | 78 (46.4) | 136 (41.5) |
| college | 79 (49.4) | 63 (37.5) | 142 (43.3) |
| Title |  |  |  |
| resident physician | 121 (75.6) | 145 (86.3) | 266 (81.1) |
| attending physician | 24 (15.0) | 14 (8.3) | 38 (11.6) |
| associate chief physician | 15 (9.4) | 9 (5.4) | 24 (7.3) |
| Working years | 14.6 ± 10.7 | 13.2 ± 9.6 | 13.9 ± 10.3 |

The trends of antibiotic prescription and inappropriate antibiotic prescription rates over time in the two groups are shown in Figure 6. The bottom half of the figure shows trends in antibiotic prescription rates for group 1 (blue triangles) and group 2 (red circles). At baseline, there was no statistically significant difference in antibiotic prescription rates between the two groups (*p* = 0.085). The rate of antibiotic prescription in group 1 decreased significantly after 20 days and then gradually leveled off. The antibiotic prescription rate of group 2 also decreased, but the change was not as obvious compared with group 1. At the end of the first period (at the cross-over point), the antibiotic prescription rate in group 1 decreased by 11.9% (*p* < 0.001) and in group 2 by 4.5% (*p* < 0.001). In period 2, after the cross-over, the rate of antibiotic prescriptions in group 1 increased intially but decreased overall (ΔAPR = 2.6%, *p* = 0.045). At this stage, group 2 was receiving the intervention and maintained their downward trend (ΔAPR = 11.7%, *p* < 0.001).

**Figure 6.**
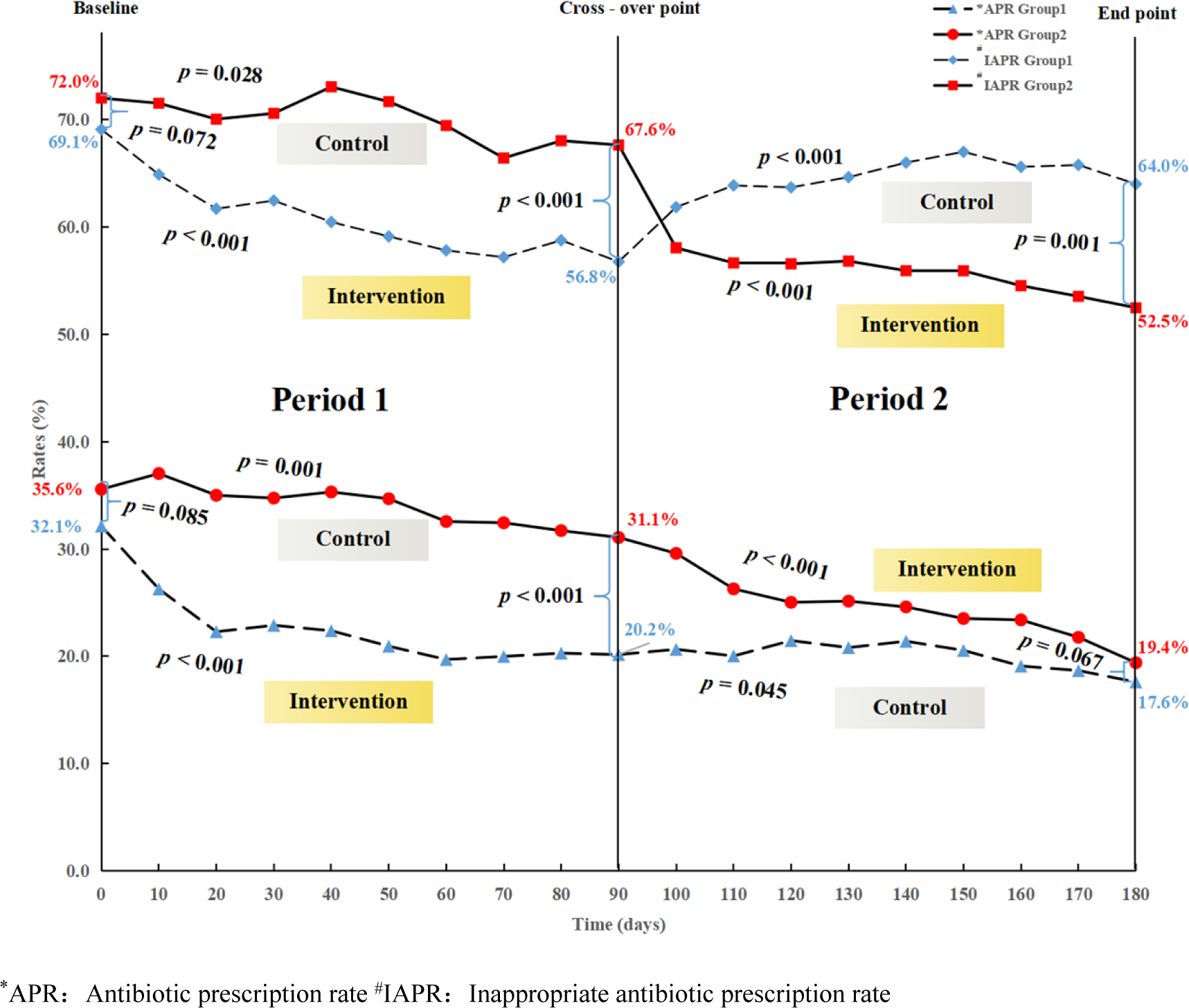
Comparison of antibiotic prescription rates and inappropriate antibiotic prescription rates over time between the two groups.

The top half of Figure 5 shows trends in inappropriate antibiotic prescription rates for group 1 and group 2. In Period 1, the rate in group 1 decreased by 12.3% (*p* < 0.001), while in group 2 the rate decreased by 3.1% (*p* = 0.028). In period 2, group 2 received the intervention, and the inappropriate antibiotic prescription rate decreased rapidly at the beginning and then gradually decreased (ΔAIR = 15.9%, *p* < 0.001). In group 1, who crossed over to the control, the inappropriate antibiotic prescription rate increased to 64.1% (*p* < 0.001) by the end of the study.

To predict changes in antibiotic prescription and inappropriate antibiotic prescription rates over time, two transition models were fit to the data with the results shown in Table 2. The coefficients represent changes in rates under the influence of explanatory variables. The intercept coefficients of antibiotic prescription rate and inappropriate rate were -0.05 (*p* = 0.005)/ -0.04 (*p* = 0.616), indicating that when all variables were controlled for in the initial state, antibiotic prescription and inappropriate antibiotic prescription rates decreased by 5% and 4% every 10 days, respectively. The respective coefficients for the feedback intervention were -0.05 (*p* < 0.001) and -0.04 (*p* = 0.046), meaning that the intervention resulted in a 5% and 4% reduction in 10-day antibiotic and inappropriate antibiotic prescribing rates. The coefficients for time point were 0.01 (*p* = 0.009) and 0.007 (*p* = 0.037), for antibiotic prescriptions and inappropriate antibiotic prescriptions, respectively and for period were -0.04 (*p* = 0.041) and -0.05 (*p* = 0.145), respectively, indicating that both rates decreased gradually with the passage of time, and the decreasing rates in period 2 were 4% and 5% lower than that of period 1. The correlation coefficients of physicians’ demographic characteristics were not significant in both models. Period was also not significant for inappropriate antibiotic prescription rates.

**Table 2.** Transition model predicting the change in antibiotic prescription rates and inappropriate antibiotic prescription rates.

| Characteristic | Coef. | 95% CI | $p$ (t-test) | $p$ (F-test) |
| --- | --- | --- | --- | --- |
| (Intercept) | -0.05 / -0.04 |  | 0.005 / 0.616 | 0.005 / 0.616 |
| Feedback (intervention vs control) | -0.05 / -0.04 | (-0.07, -0.03) / (-0.07, 0) | < 0.001 / 0.046 | < 0.001 / 0.046 |
| Time point (1–19) | 0.01 / 0.007 | (0.00, 0.01) / (0, 0.01) | 0.009 / 0.037 | 0.009 / 0.037 |
| Period (2 vs 1) | -0.04 / -0.05 | (-0.08, 0.00) / (-0.13, 0.02) | 0.041 / 0.145 | 0.041 / 0.145 |
| Physicians' characteristic |  |  |  |  |
| Sex: male vs female | 0.00 / 0.00 | (-0.02, 0.02) / (-0.04, 0.04) | 0.872 / 0.917 | 0.872 / 0.917 |
| Age: ref. = 22–31 years |  |  |  | 0.992 / 0.999 |
| 32–41 | 0.00 / 0.00 | (-0.03, 0.03) / (-0.05, 0.05) | 0.908 / 0.991 |  |
| 42–65 | 0.00 / 0.00 | (-0.07, 0.06) / (-0.11, 0.11) | 0.908 / 0.968 |  |
| Education: ref. = college |  |  |  | 0.919 / 0.942 |
| Junior college | 0.00 / 0.00 | (-0.03, 0.02) / (-0.04, 0.04) | 0.687 / 0.95 |  |
| Technical secondary school | 0.00 / 0.01 | (-0.04, 0.03) / (-0.05, 0.07) | 0.815 / 0.774 |  |
| Title: ref. = Associate chief physician |  |  |  | 0.99 / 0.996 |
| Attending physician | 0.00 / 0.00 | (-0.05, 0.06) / (-0.09, 0.09) | 0.91 / 0.98 |  |
| Resident physician | 0.00 / 0.00 | (-0.04, 0.05) / (-0.08, 0.08) | 0.886 / 0.972 |  |
| Working years | 0.00 / 0.00 | (0.00, 0.00) / (-0.01, 0) | 0.947 / 0.877 | 0.947 / 0.877 |

\*APR: Antibiotic prescription rate
#IAPR: Inappropriate antibiotic prescription rate

Figure 7 shows the correlation of antibiotic prescription rate and inappropriate antibiotic prescription rate between the two groups under the intervention state. The correlation coefficients are 0.954 (*p* < 0.001) and 0.947 (*p* < 0.001), respectively.

**Figure 7.**
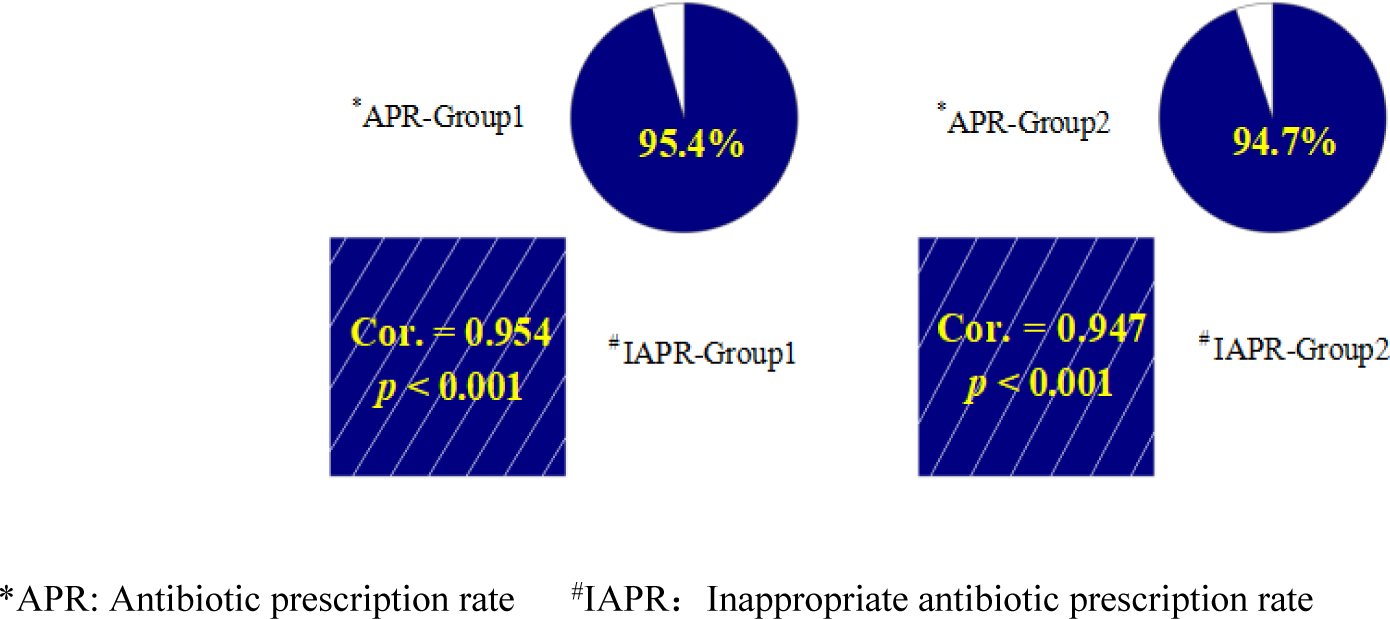
Correlations between antibiotic prescription rates and inappropriate antibiotic prescription rates under intervention condition. **Notes:** In Figure 7, the correlations between the antibiotic prescription rate and inappropriate antibiotic prescription rate are shown. The box in the lower left corner indicates the relevant direction; blue means positive. The top right pie chart shows the strength of the correlation. The larger areas of the color portion of the pie, the stronger the correlation.

Figure 8 shows a strong negative correlation between antibiotic prescription ranking and number of clicks on the pop-up during the intervention period (r = 0.591, p < 0.001). The lower the physicians’ rankings (representing lower rates of antibiotic prescriptions), the higher the number of clicks.

**Figure 8.**
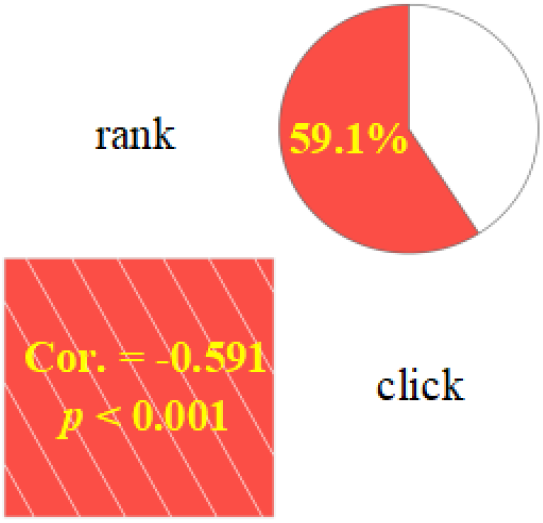
Correlation between antibiotic prescription ranking and number of clicks under intervention conditions. **Notes:** In Figure 8, the correlation between antibiotic prescription rate ranking and number of clicks during the intervention period are shown. The box in the lower left corner indicates the relevant direction; red means negative. The top right pie chart shows the strength of the correlation. The larger areas of the color portion of the pie, the stronger the correlation.

## Discussion

In this study, we attempted to build on the previous study [23] with a more comprehensive feedback intervention in new primary care institutions (n=77) and outpatient physicians (n=328). These interventions aimed to provide antibiotic prescription feedback to physicians every 10 days, real-time warnings of inappropriate antibiotic prescriptions, and educational brochures. The latter two interventions were added from our previous study [23]. These interventions were significant and stable in reducing antibiotic prescription rates by outpatient physicians in primary care institutions. In terms of the inappropriate antibiotic prescription rate, the effect was evident in the first intervention period but had an upward trend in the second control period. None of the physician characteristics were associated with antibiotic prescription or inappropriate prescription rates.

From baseline to endpoint, the antibiotic prescription rates of the two groups were reduced by 14.5% (group 1) and 16.2% (group 2) This is close to the reduction in antibiotic prescription rates seen in our previous study [23]. Recent studies have shown that interventions can reduce antibiotic prescription rates by 5.6% to 22% [15, 38, 39]. However, the limitation of our previous study [23] was that it only considered the reduction of antibiotic prescription rate, without considering the rationality of antibiotics. As a result, the primary care institutions and physicians focused only on reducing antibiotic prescriptions, regardless of whether antibiotic prescriptions were justified. Therefore, an effective intervention can only be achieved if both the overall prescribing rate and the inappropriate prescribing rate of antibiotics are reasonably reduced. In addition, we also collected secondary diagnostic information in this study. Therefore, the judgment of inappropriate antibiotic prescription rates was more reasonable.

The feedback intervention measures mainly reminded the outpatient physicians to pay attention to their prescribing behavior through real-time early warning feedback and information feedback once every 10 days. Real-time warning pop-up messages are used to intervene when the physicians are about to make an inappropriate prescribing behavior. This intervention makes them immediately aware that there might be a problem with their prescription. Physicians can choose to change prescriptions or ignore these warnings. The intervention is not mandatory, which may be why it was widely supported by physicians and primary care institution leaders during the study period. In addition, the confidentiality and non-coercive nature of the 10-day antibiotic prescription feedback, which continues the previous study [23], made it more acceptable to the physicians. Moreover, in this study we collected the number of mouse clicks made by physicians looking at information about prescriptions. There was a negative correlation between the number of clicks and antibiotic prescription rate. This indicates that the physicians with high compliance were more likely to change their prescribing behavior. Future researchers should devise more interventions in which physicians voluntarily participate. In addition, the educational brochures distributed were introduced in our intervention protocol [29]. We developed the educational materials specifically for primary care physicians using the Delphi method [40]. Distribution of educational manuals is a common educational intervention and does not fall within the scope of a feedback intervention [18, 31]. However, it provides a powerful aid for our feedback intervention. In the pre-intervention research phase, the physicians liked this brochure, which was provided free of charge. It was conducive to the further development of feedback intervention measures.

In the intervention period, the antibiotic and inappropriate antibiotic prescription rates of both groups showed a rapid decline, followed by fluctuations. This phenomenon may be explained by the transtheoretical model [41], which interprets behavior change as a continuous, dynamic, and gradual process [42]. It is divided into 5 stages, including precontemplation, contemplation, preparation, action and maintenance, and the transformation of each stage requires a process [43]. Real-time warning pop-up messages accelerate the first four stages of the process enabling physicians to consider behavior changes and act in just a few minutes. The contamination effect and Hawthorne effect also explain why the first four stages of the transtheoretical model were so rapid in our study. As a result, antibiotic prescription rates in the intervention group initially declined rapidly. However, due to obstacles such as physician prescribing habits, patient interference, and distrust by some physicians, not everyone can complete all five stages [44, 45]. This is probably why there were two slight rises in the prescription rates during the intervention period. In the end, the intervention effect was consolidated again due to the repeated reinforcement of prescription information reminders and educational brochure given once every 10 days, and the overall change of physicians’ prescribing behavior was finally realized.

We observed a rebound in inappropriate antibiotic prescription rates after the transition from intervention to control. This may be due to the generally low professional and technical level of physicians in primary care institutions in China [46, 47]. Without the real-time warning pop-up message, physicians would not have been able to realize when they were prescribing inappropriate antibiotics. Due to the delayed effect of behavioral interventions [48], they might blindly reduce antibiotic prescriptions even in the absence of an intervention, but could not effectively change their inappropriate prescription behavior. This result may also reflect the high compliance of physicians to the real-time warning pop-up message. But overall, from our correlation test, there was a strong positive correlation between the rate of antibiotic prescription and the rate of inappropriate antibiotic prescription under the intervention conditions.

To further explore the effects of the intervention, transition models were used to predict the impact of antibiotic and inappropriate antibiotic prescription rates. As expected, the effect of the feedback intervention was significant and stable, which might be attributed to the real-time warning pop-up message and educational brochures provided to the physicians. Further validation of the relationship in the inappropriate antibiotic prescription rates between two periods may be required.

In conclusion, the combination of real-time warning pop-up messages, antibiotic prescription feedback, and educational brochures enabled primary care physicians to prescribe antibiotics appropriately and effectively reduce their antibiotic prescription rates. Our study plays an important role in reducing the risk of antibiotic resistance. This new intervention may be preferable to our previous study [23].

There are some limitations to the current study. Firstly, it is difficult to prevent communication between physicians in different primary care institutions. Physicians in the control group of one institution can inform their colleagues in other institutions of the intervention content in advance, resulting in a contamination effect [49]. Secondly, all antibiotic prescribing data in our study came from the HIS. Primary care institutions often run out of antibiotics. Some physicians gave their patients paper prescriptions and instructed them to visit the pharmacy to buy the prescribed antibiotics. As a result, the actual number of antibiotic prescriptions may be higher than what was reported in this study.

## Data Availability

All relevant data are within the manuscript and its Supporting Information files.

## Acknowledgments

We thank all the participating institutions for providing information and assistance during the study. The authors also thank all members of the investigational team who collected the data. We acknowledge the assistance of Edward McNeil from The Chinese University of Hong Kong for proof-reading the manuscript and providing suggestions to improve it.

## Declaration

### Consent for publication

Not applicable.

### Availability of data and materials

The datasets used and/or analysed during the current study are available from the corresponding author on reasonable request.

## Authors’ contributions

Conceptualization: YC and ZZC;

Data curation: STY;

Formal analysis: YC and JLY;

Funding acquisition: YC;

Investigation: YC, JLY, WD and SYW;

Methodology: YC and JLY;

Project administration: YC;

Supervision: XJL and XH;

Resources: XJL and XH;

Software: YC and JLY;

Visualization: YC and ZZC;

Writing – original draft: JLY;

Writing – review & editing: YC and ZZC

## Abbreviations

CDSS: Clinical Decision Support System
HIS: Hospital Information System
DGNN: Depth Graph Neural Network technology
AI: artificial intelligence
LWTC: Lianke Weixin Technology Co., LTD.。
ICGPHC: Information Center Guizhou Provincial Health Commission
ICD-10: International Classification of Diseases 10^th^ Edition
APR: Antibiotic prescription rate
IAPR: Inappropriate antibiotic prescription rate

## Supporting information

S1 Appendix 1 CONSORT 2010 checklist of information to include when reporting a cluster randomized trial

S2 Appendix 2 Certificate of approval for ethical review

S3 Appendix 3 The brochures cover, catalogue, and example of content

S4 Appendix 4 Catalogue for Clinical Application of Antibacterial Drugs

S5 Appendix 5 Metadata

